# Clarifying the anatomy of tetralogy of Fallot with S-shaped ascending aorta

**DOI:** 10.1101/2024.01.04.24300875

**Authors:** Saurabh Kumar Gupta, Aprateem Mukharjee, Niraj Nirmal Pandey, Sivasubramanian Ramakrishnan, Shyam Sunder Kothari, Anita Saxena, Robert H Anderson

## Abstract

**Background:** We have recently encountered several cases of tetralogy of Fallot with an abnormally oriented S-shaped ascending aorta. In this retrospective study, we sought to clarify the morphology of this unusual under-recognized variant of tetralogy of Fallot.

**Methods:** We reviewed our databases to identify all patients with tetralogy of Fallot having an S-shaped ascending aorta. We then used computed tomographic angiography to make a detailed assessment of the cardiac morphology.

**Results:** Out of the 21 patients we identified, 18 (86%) had a right aortic arch, 2 (9%) had a left aortic arch, and the remaining patient (5%) had a double aortic arch. Patients with right aortic arch, compared to those with the normally oriented ascending aorta, had lesser aortic override (29.3±14% vs 54.8±13.2%; p=0.0001) and a wider ascending aorta (2.52±0.7 cm vs 1.80±0.32; p=0.0003). Overall, compared to normal cases, the ascending aorta was located posteriorly, with a higher sterno-aortic distance (2.55±0.8 cm vs 0.99±0.45 cm; p=0.0001). The ascending aorta was longer (4.12±1.7 vs 3.07±0.82, p=0.03) although the tortuosity index (1.22±0.19 vs 1.15±0.17, p=0.23) was not different. Of the cases with right aortic arch and S-shaped ascending aorta, 16 (89%) had extrinsic compression of the right pulmonary artery (p = 0.0001), while 7 (39%) had crossed pulmonary <u>arteries</u> (p = 0.008), with no such findings in those with normally oriented ascending aorta or those with left aortic arch and S-shaped ascending aorta.

**Conclusion:** Tetralogy of Fallot with an S-shaped ascending aorta is a variant with lesser aortic override and a more posterior location of the aorta. Compression of the right pulmonary artery and crossed pulmonary arteries are frequent when the arrangement is associated with a right-sided aortic arch. These findings may have important implications for surgical management.

## Introduction

Tetralogy of Fallot is one of the most common congenital heart diseases causing arterial desaturation. Its phenotypic feature is anterocephalad deviation of the outlet septum coupled with abnormal septoparietal trabeculations, thus producing a malaligned ventricular septal defect and right ventricular outflow tract obstruction. (1–3) While the dimensions of the pulmonary arteries are the main determinant for surgical repair, other morphological factors also alter the surgical technique and outcomes. Besides its phenotypic features, there is a wide spectrum of associated morphological abnormalities (1–5). Some of these may be benign variations, whereas others have significant implications for surgical management. While echocardiography remains the main diagnostic imaging modality, other imaging modalities, such as computed tomographic angiography, catheter angiography, or magnetic resonance imaging, are often performed for preoperative assessment (5). Computed tomographic angiography in particular, owing to its high spatial resolution, provides excellent visualization of both the cardiac and extracardiac anatomy (6–9).

Some time ago, we reported a patient whose right pulmonary artery was unusually compressed by a tortuous aorta. In this patient, the left pulmonary artery was not arising from the pulmonary trunk but instead was supplied by a restrictive patent arterial duct (10). Unlike other cases with a right aortic arch, despite its usual origin from the heart, the ascending aorta was tortuous, with its proximal part lying unusually leftward and posteriorly. The child successfully underwent surgical repair with a transannular patch and reimplantation of the left pulmonary artery. Aortic transection and the Lecompte maneuver were deemed necessary to relieve extrinsic compression of the right pulmonary artery (10–12). Since then, we have encountered several additional cases with abnormally oriented and tortuous ascending aorta, the majority having extrinsic compression of the mid-segment of the right pulmonary artery. (13) A detailed review of the literature indicated a paucity of information about this particular variant. In this retrospective study, therefore, we sought to clarify the morphology of this unusual variant of tetralogy of Fallot with S-shaped ascending aorta, using computed tomographic angiography as the main imaging modality.

## Methods

Following approval from the institutional ethics committee, we searched our databases from January 2012 to October 2023 to identify all patients with tetralogy of Fallot and abnormally oriented ascending aorta. The demographic details were obtained from medical records. All patients had had echocardiographic evaluation by a pediatric cardiologist, and computed tomographic evaluation by a cardiac radiologist. In select cases, cardiac catheterization studies had also been performed by a pediatric cardiologist. Computed tomographic studies were analyzed using a Syngovia workstation (Siemens Healthcare, Germany). All the imaging data were retrospectively analyzed independently by a pediatric cardiologist and a cardiac radiologist. The differences were resolved by consensus, obtaining inputs from other pediatric cardiologists and cardiac radiologists. Besides the identified cases, we also analyzed computed tomographic angiograms of age and sex-matched patients having tetralogy of Fallot with normally oriented ascending aorta. Since the majority of patients with abnormally oriented ascending aorta had a right aortic arch, the comparison was made only for those with a right aortic arch. For each case, we included one age and sex-matched control. After evaluating the segmental cardiac anatomy, we analyzed all the morphological variables in a predefined fashion, placing special emphasis on the spatial relationship of the arterial valves, the orientation of great arteries, the arrangement of the right and left pulmonary arteries, and the sidedness of the aortic arch. We also noted the presence of crossing of the pulmonary arteries, the origin and course of coronary arteries, and any anomalies of the aortic arch. The analysis of the aortic root and aorta was performed after obtaining the true short axis of the aortic root using the double oblique technique. (14) After a multiplanar assessment, all measurements were made in two orthogonal planes. The analysis was supplemented by volume rendering and virtual dissection (9).

The sterno-aortic and sterno-vertebral distances were measured to define the relative position of the ascending aorta in the mediastinum. The distance between the ascending aorta and sternum was taken midway between the sinutubular junction and the origin of the first branch of the aorta (Figure 1A). The length of the ascending aorta was assessed by measuring its length in the centreline, as well as the shortest length between the sinutubular junction and the first brachiocephalic artery. We then calculated the tortuosity index, defined as the ratio between the centreline distance and the shortest length of ascending aorta (Figure 1B). (15)

**Figure 1:**
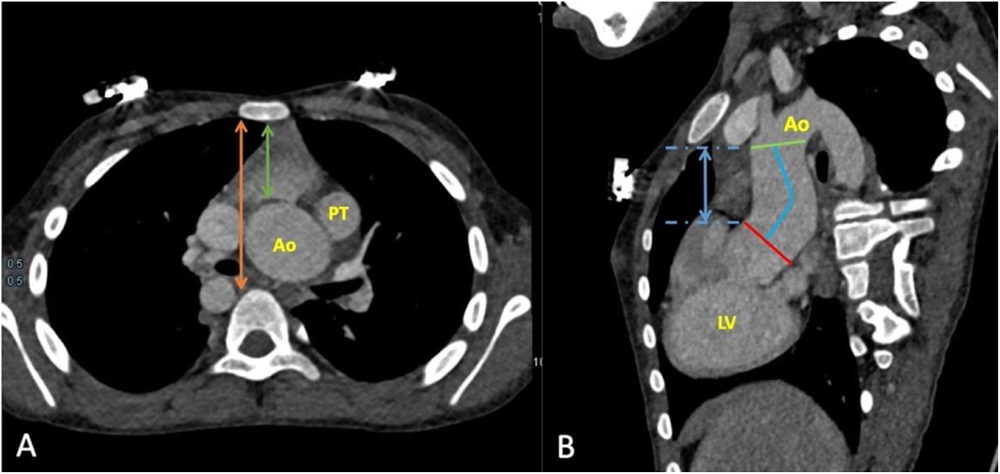
Computed tomographic angiography images in axial (A) and oblique sagittal planes (B) show the posteriorly located and tortuous ascending aorta. Panel A shows the technique used for the measurement of the sterno-aortic distance (green double arrow) and the sterno-vertebral distance (orange double arrow) midway between the sinutubular junction and the origin of the first branch of the aorta. Panel B shows the tortuosity index which was calculated as the ratio of the centreline distance (blue curved line) and the shortest length (blue double arrows) between the sinutubular junction and the first brachiocephalic artery.

### Statistical analysis

Categorical data were represented by percentages or frequencies. They were compared using Fisher’s exact test. Continuous variables were summarized as median with range, or mean with standard deviations, and were compared using an independent t-test. A p-value of <0.05 was considered statistically significant. All statistical analyses were performed using IBM SPSS V.23 software.

## Results

We identified 21 patients with abnormally oriented ascending aorta. The age of the patients ranged from 2 months to 21 years. Their oxygen saturation ranged from 65 to 85%. All had normal sequential segmental anatomy. In all patients, there was a solitary large malaligned ventricular septal defect and an obstructed right ventricular outflow tract (Figures 2 and 3; Videos 1-3). All except one patient had fibrous continuity between the leaflets of the aortic and mitral valves.

**Figure 2:**
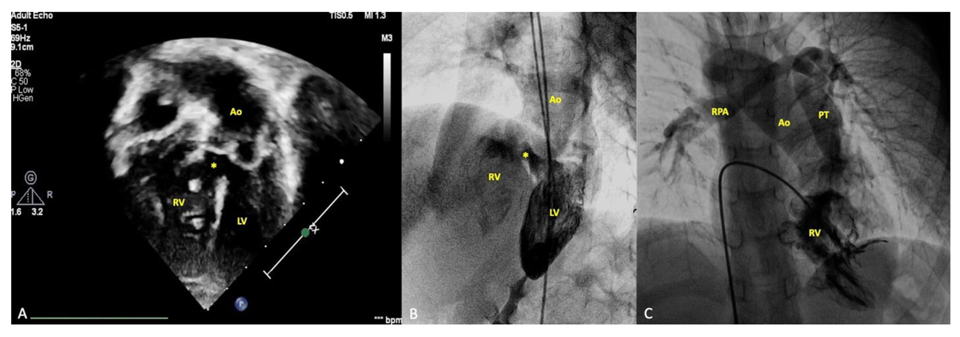
Echocardiogram in subcostal view (A) and catheter angiogram in anteroposterior cranial (B) and left anterior oblique (C) views show a malaligned ventricular septal defect (*) characteristic of tetralogy of Fallot but with less than 50% aortic override and a posteriorly located tortuous ascending aorta. Right aortic arch, crossed pulmonary arteries and right pulmonary artery stenosis are also evident in panel C. These findings are better appreciated in videos 1-3.

**Figure 3:**
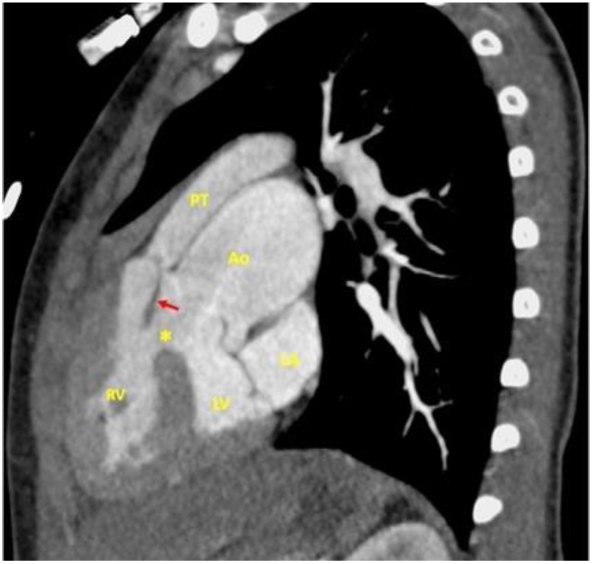
Computed tomographic angiography image in the sagittal plane shows malaligned ventricular septal defect (*) and anterocephalad deviation of the outlet ventricular septum (red arrow) characteristic of tetralogy of Fallot. The aortic override, however, is less than 50% with the aorta mostly committed to the left ventricle.

### Ventricular outflows, aortic override, arterial valves and arterial trunks

In all cases, the ventricular outflow tracts were normally oriented, with aortic override ranging from 10 to 45%. The override was significantly less in cases with a right-sided aortic arch compared to controls having a normal orientation of the ascending aorta (29.3±14% vs 54.8±13.2%, p 0.0001) (Figures 2 and 3; Videos 1-3).

One patient had pulmonary atresia. In the remaining 20 patients with pulmonary stenosis, the pulmonary valve was superior to the aortic valve, being to the left and anterior of the aortic valve in 17 patients (88%). In 1 (5%) patient, the arterial valves were side by side, with the aortic valve lying to the right of the pulmonary valve. In the other 2 (10%) patients, the pulmonary valve was located right and anterior to the aortic valve. In most patients, we noted that the subpulmonary outflow tract was poorly visualized on transthoracic echocardiography.

### Ascending aortic length, orientation, and tortuosity

In all patients, the ascending aorta was tortuous, producing an S-shaped orientation when viewed from the head end (Figures 1 and 4 and Video 4). The tortuosity was less pronounced among patients with left-sided and double aortic arches. Compared to controls, those with right-sided aortic arch had longer ascending aorta (41.2±17.0 mm vs 30.7±8.2 mm, p 0.025), although the tortuosity index, as well as the shortest distance between the sinutubular junction and the first aortic branch, were not different (Table 1). The ascending aorta within the overall group was also wider when compared to controls, being the widest above the sinutubular junction (25.2±6.9 mm vs 18.0±3.2 mm, p 0.0003). Among those with tortuous aorta, the ascending aorta was located more posteriorly, with a greater sterno-aortic distance (25.5±7.7 mm vs 9.9±4.5 mm; p 0.0001), but with a similar sterno-vertebral distance (58.1±14.1 vs 51.5±6.6, p 0.08) (table 1).

**Figure 4:**
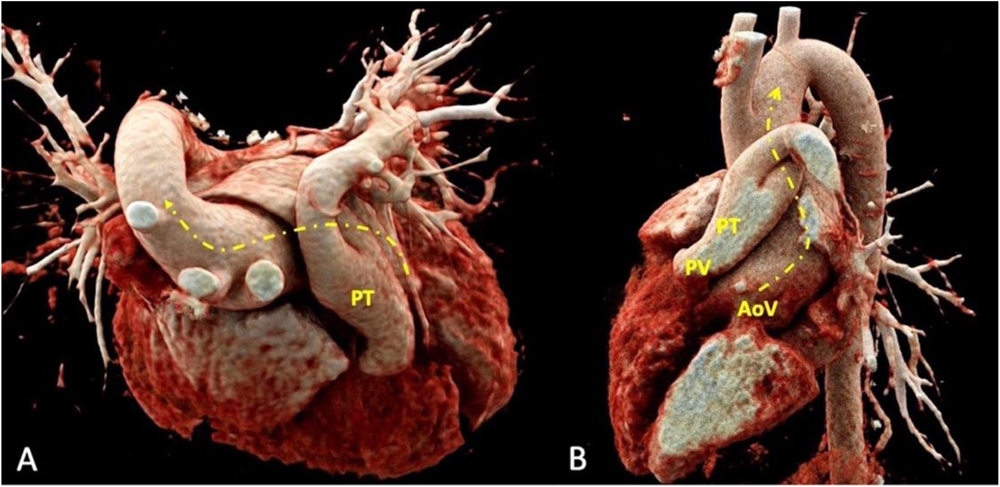
Volume-rendered images seen from the head end (A) and in the left anterior oblique view show the tortuous S-shaped ascending aorta (dashed arrows in A and B). Crossed pulmonary arteries, less than 50% aortic override, and a narrowed mid-segment of the right pulmonary artery are also evident. AoV – Aortic valve; PT – Pulmonary trunk; PV - Pulmonary valve

**Table 1:**
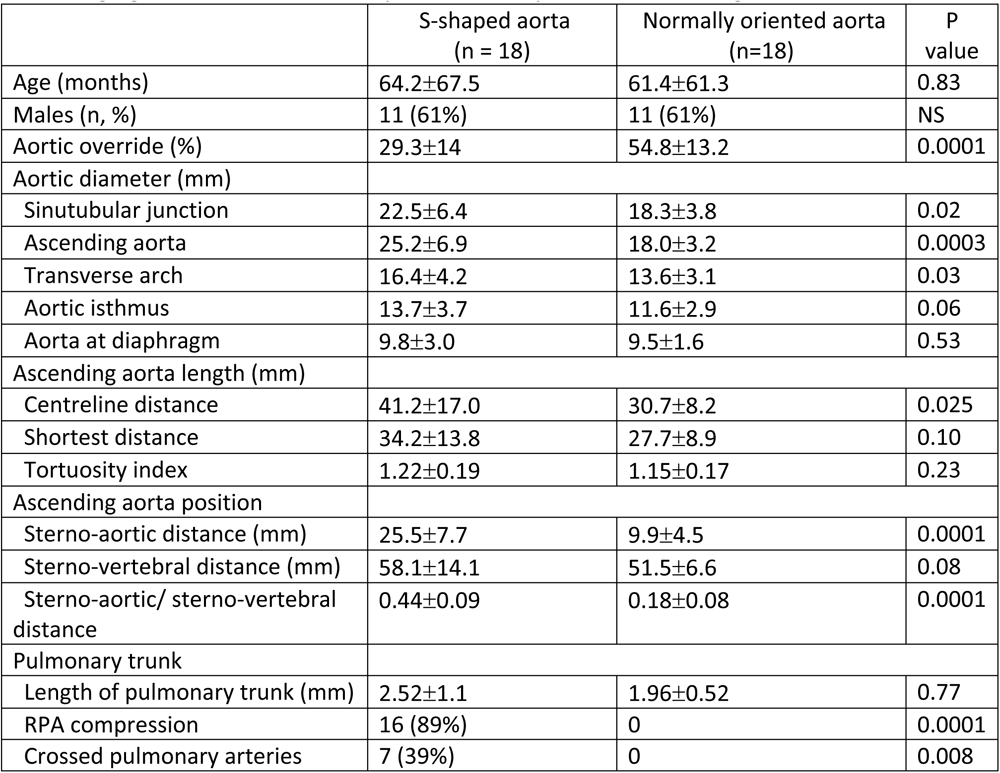
Demographic and morphologic characteristics of patients with tetralogy of Fallot having right aortic arch with S-shaped or normally oriented ascending aorta.

### Right ventricular outflow tract and pulmonary arteries

The right ventricular outflow tract was obstructed in all patients. It was atretic in one patient, with all the remainder having subpulmonary stenosis. The length of the pulmonary trunk was not different from the controls (Table 1). Of the 18 cases with right aortic arch, 16 (89%) had compression of the mid-segment of the right pulmonary artery. No such compression was found in the remaining 5 (28%) patients. Of these patients, 2 had a right arch, 2 a left arch, and the remaining a double arch. In those with compression of the right pulmonary artery, it involved the mid-segment near the right bronchus. The compression in the anteroposterior direction gave the right pulmonary artery an oval shape (Figures 2C and 5 and Video 5). In addition, 3 (21%) of our patients, and 1 control, also had stenosis or atresia of the orifice of the left pulmonary artery. In 7 of our 18 (39%) cases with right arch, the pulmonary arteries were crossed, a feature not found either in the controls or in the patients with left or double aortic arch (Figures 2C and 5C).

**Figure 5:**
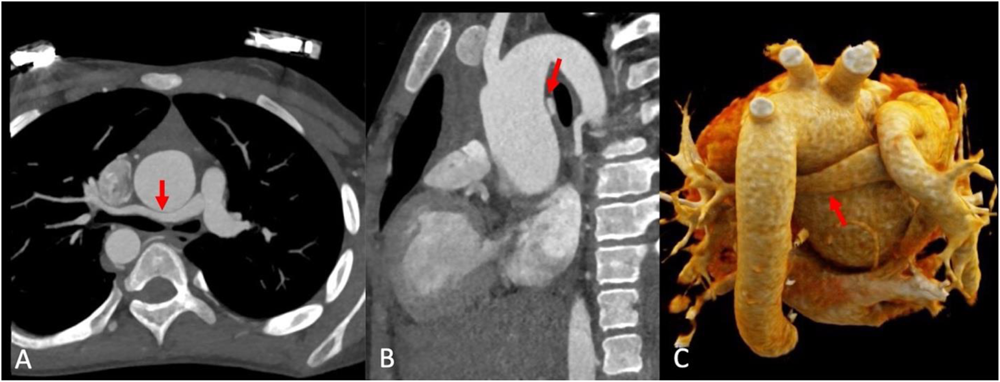
Computed tomographic angiographic images in axial (A) and sagittal (B) views, and a volume-rendered 3D reconstruction (C), show compression of the mid-segment of the right pulmonary artery between the dilated tortuous ascending aorta anteriorly and right-sided descending thoracic aorta posteriorly. Crossed pulmonary arteries are also evident in panel C.

### Branching of the aortic arch

The aortic arch was right-sided in 18 (86%) patients, being left-sided in 2 (9%), and double in the other (5%). In all except 3 patients, the branching pattern of the brachiocephalic vessels was as expected for the side of the aortic arch. The patient with a double aortic arch had the left common carotid and left subclavian arteries arising from the left arch, while the comparable right-sided vessels arose from the right arch. Another patient with a right-sided aortic arch had isolation of the left subclavian artery, while 1 patient had an aberrant retroesophageal left subclavian artery.

### Coronary arteries

The coronary arteries arose in a normal fashion in all but one patient. In the outstanding patient, who had a right-sided aortic arch, the left circumflex coronary artery arose from the right coronary artery taking a retro-aortic course.

## Discussion

Tetralogy of Fallot is immediately recognizable based on a malaligned ventricular septal defect and characteristic infundibular phenotype. Morphologic variations are well recognized. It seems, however, that the variant with an S-shaped aorta has thus far escaped attention. We have now discovered this unusual finding in 21 of our patients. The tortuous aorta is part of a relatively uniform constellation of abnormalities. Not only is the ascending aorta dilated and tortuous, but it is also located relatively posteriorly. In those with right-sided aortic arch, this results in overcrowding of vascular structures in the posterior mediastinum, producing anteroposterior compression of the mid-segment of the right pulmonary artery between the ascending aorta anteriorly, and the right bronchus and the right-sided descending aorta posteriorly. In contrast, despite similar tortuosity of the ascending aorta, compression of the right pulmonary artery was lacking in those with left-sided and double arches. The older age of patients possibly explains our finding of frequent compression of the right pulmonary artery as aortic dilation is known to be progressive and age-dependent in tetralogy of Fallot. Also, aortic dilation is inversely proportional to the flow of blood through the pulmonary arteries. Those with less pulmonary flow tend to have larger aorta and smaller right pulmonary artery, leading to an increased likelihood of compression. Recognition of the extrinsic compression as the cause of the narrowing of the right pulmonary artery is important in these patients. As in the treatment of our first case, (10) the Lecompte maneuver, or aortopexy, may be needed to achieve sustained relief from the extrinsic compression produced by the dilated ascending aorta (16).

Crossing of the pulmonary arteries is also unusual in patients with tetralogy of Fallot (17). More than one-third of our patients with right aortic arch, nonetheless, had crossed pulmonary arteries. We speculate that the coexistence of this finding with the tortuous ascending aorta may indicate abnormal rotation of the developing proximal outflow tract and arterial trunks as the underlying mechanism producing tetralogy of Fallot with S-shaped ascending aorta. The aortic override was much less in our patients than in their controls. This, again, is unlike the usual situation with tetralogy of Fallot. In an autopsy study, the aortic override was found to range from 31 to 100%, with most individuals having greater than 50% override. (18) In our study, all of our patients had an override of less than 45%. (18) An abnormal location of the aortic valve relative to the pulmonary valve, furthermore, was observed in only 3% of the autopsy specimens. (18) The finding of an abnormally located aortic valve in one-tenth of our cohort likely reflects the selective inclusion of patients with tortuous ascending aorta.

We were perplexed to find such a large number of patients with a seemingly unrecognized variant of tetralogy of Fallot. While making a more detailed literature review, we found that a tortuous ascending aorta is also part of the unusual arrangement variously described as ‘anatomically corrected malposition of great arteries’, ‘isolated infundibuloarterial inversion’, or ‘TOF {S,D,I}’. (19–23). The additional feature proposed for these cases, however, is that the arterial trunks extend in side-by-side fashion into the mediastinum, rather than spiralling as was the situation in our cases. While an unusually lesser degree of override, combined with the more posterior location of the aortic valve, gives an appearance of side-by-side arterial trunks, the overall anatomy remains that of tetralogy of Fallot. Moreover, since the surgical management is similar to tetralogy of Fallot, we believe that it is better to describe details of the overall arrangement of the ventricular outflows and arterial trunks in such situations (24,25). It is arguably the use of ill-defined terminologies, such as “infundibulo-arterial inversion”, and the lack of a systematic approach, which has left the S-shaped tortuous ascending aorta as an unrecognized entity, despite its unique morphologic characteristics. One of the cases previously reported by one of us has all the features we have now encountered in our overall series. On a detailed review, we realized that the case has no specific morphologic characteristics to justify the previously chosen label of isolated “infundibulo-arterial inversion”. (13) Our ongoing experience exemplifies the difficulties that have previously been encountered in understanding ill-defined variants of common cardiac malformations.

Several of the previously described examples of tetralogy of Fallot with alleged “infundibulo-arterial inversion” have striking similarities to our cases. (13,16,21). Not only was the ascending aorta tortuous in some of these but the pulmonary arteries were also crossed, just as we have observed (13,16). Compression of the right pulmonary artery was not seen in the case reported by Nelson and colleagues, which had side-by-side arterial trunks, but there was extrinsic compression of the distal right bronchus (16). The authors attributed the finding to a low-lying dilated transverse arch. It is possible, nonetheless, that the mechanism could be the same as in our cases. The lack of compression of the right pulmonary artery in their case may well be related to the young age of the patient.

Based on the findings of our study, we suggest that a finding of less than 50% aortic override, and any suggestion of tortuosity of the aorta during the echocardiographic study, should immediately raise the suspicion of an S-shaped ascending aorta. Difficulty in visualizing the subpulmonary infundibulum during echocardiography could also provide a clue towards the diagnosis. This should also prompt a detailed assessment of the right pulmonary artery for any extrinsic compression. With our own increased experience, we have now been able to identify many of our patients confidently using echocardiography, although computed tomographic angiography was performed to delineate the morphologic findings to optimize the planned surgical intervention.

We recognize that our study is limited by its retrospective nature and that our cohort is still relatively small. We also recognize a possible association with DiGeorge syndrome, but genetic testing could not be performed. Only a few patients underwent surgery, so surgical correlation was not possible. We conclude, nonetheless, that a tortuous and S-shaped ascending aorta is a discrete but previously unrecognized morphologic variant of tetralogy of Fallot, with clinical and surgical implications. We hope our findings might help others in the prompt detection and appropriate surgical planning of this unusual variant.

## Data Availability

All data would be made available on request.

## References

1. Evans WN. “Tetralogy of Fallot” and Etienne-Louis Arthur Fallot. Pediatr Cardiol 2008;29:637–40.

2. Anderson RH, Moorman A, Brown N, Bamforth S, Chaudhry B, Henderson D et al. Normal and abnormal development of the heart. In: Da Cruz E, Ivy D, Jaggers J (eds). Pediatric and Congenital Cardiology, Cardiac Surgery and Intensive Care. London: Springer, 2013, 151–77.

3. Anderson RH, Jacobs ML. The anatomy of tetralogy of Fallot with pulmonary stenosis. Cardiol Young 2008;18:12–21.

4. Van Praagh R. The first Stella van Praagh memorial lecture: the history and anatomy of tetralogy of Fallot. Semin Thorac Cardiovasc Surg Pediatr Card Surg Annu 2009;12:19– 38.

5. Anderson RH, Spicer DE, Henry GW, Rigsby C, Hlavacek AM, Mohun TJ. What is aortic override? Cardiol Young 2015;25:612–25.

6. Chelliah A, Shah AM, Farooqi KM, Einstein AJ, Han BK. Cardiovascular CT in cyanotic congenital heart disease. Curr Cardiovasc Imaging Rep 2019;12:30

7. Siripornpitak S, Pornkul R, Khowsathit P, Layangool T, Promphan W, Pongpanich B. Cardiac CT angiography in children with congenital heart disease. Eur J Radiol. 2013;82:1067–82.

8. Sachdeva S, Gupta SK. Imaging modalities in congenital heart disease. Ind J Pediatr 2020;87:385–97.

9. Gupta SK, Spicer DE, Anderson RH. A new low-cost method of virtual cardiac dissection of computed tomographic datasets. Ann Pediatr Cardiol. 2019;12:110–6.

10. Talwar S, Gupta SK, Muthukkumaran S, Murugan MK, Airan B. Unusual compression of the right pulmonary artery by the aortic arch. Ann Thorac Surg 2014;97:1790–2

11. Lecompte Y, Zannini L, Hazan E, Jarreau MM, Bex JP, Tu TV, Nevux J. Anatomic correction of transposition of the great arteries. J Thorac Cardiovasc Surg 1981;82:629–31

12. Talwar S, Muthukkumaran S, Choudhary SK, Airan B. The expanding indications for the Lecompte maneuver. World J Pediatr Congenit Heart Surg 2014;5:291–6

13. Pandey NN, Sharma A, Sinha M. Isolated infundibuloarterial inversion with crisscross pulmonary arteries. Asian Cardiovasc Thorac Ann 2020;28:129–30.

14. Asch FM, Yuriditsky E, Prakash SK, Roman MJ, Weinsaft JW, Weissman G et al. The need for standardized methods for measuring the aorta: multimodality core lab experience from the GenTAC registry. J Am Coll Cardiol Imag 2016;9:219–26.

15. Ciurica S, Lopez-Sublet M, Loeys BL, Radhouani I, Natarajan N, Vikkula M, et al. Arterial tortuosity – novel implications for an old phenotype. Hypertension 2019;73:951–960.

16. Nelson JA, Soriano BD, Buddhe S. An unusual case of concordant ventriculoarterial connections, subpulmonary infundibulum, and parallel arterial trunks: a diagnostic challenge. Cardiol Young 2019;29:980–2.

17. Agarwal A, Vimalarani A, Al Amer SR, Kalis NN. Crossed pulmonary arteries: a literature review. J Ind Acad Echocardiogr Cardiovasc Imaging 2021;5:207–10

18. Khan SM, Drury NE, Stickley J, Barron DJ, Brawn WJ, Jones TJ, Anderson RH, Crucean A. Tetralogy of Fallot: morphological variations and implications for surgical repair. Eur J Cardiothorac Surg 2019;56:101–9.

19. Foran RB, Belcourt C, Nanton MA, Weinberg AG, Liebman J, Casteneda AR, Praagh R. Isolated infundibuloarterial inversion {S,D,I}: a newly recognized form of congenital heart disease. Am Jeart J 1988;116:1337–50.

20. Frank LH, Kumar TKS, Jonas RA, Donofrio MT. Tetralogy of Fallot with inverted great arteries {S,D,I}: case report, literature review, and discussion of embryology. Pediatr Cardiol 2012;33:150–4.

21. Lee ML, Chiu IS, Fang W, Chen SJ, Wang YM, Chaou WT. Isolated infundibuloarterial inversion and fifth arch in an infant: a newly recognized cardiovascular phenotypes with chromosome 22q11 deletion. Int J Cardiol 1999;71:89–91

22. Liske MR, Kavanaugh-McHugh AL, Parra DA. Isolated infundibuloarterial inversion. Pediatr Cardiol 2006;27:289–92.

23. Van Praagh R, Van Praagh S. Anatomically corrected malposition of great arteries. Br Heart J 1967;29:112–9.

24. Bernasconi A, Cavalle-Garrido T, Perrin DG, Anderson RH. What is anatomically corrected malposition? Cardiol Young 2007;17:26–34.

25. Gupta SK, Ramakrishnan S, Gulati GS, Henry GW, Spicer DE, Backer CL, Anderson RH. Clarifying the anatomy of hearts with concordant ventriculoarterial connections but abnormally related arterial trunks. Cardiol Young 2016;26:1–18.

